# Stool banking for fecal microbiota transplantation: methods and operations at a large stool bank

**DOI:** 10.1101/2020.09.03.20187583

**Authors:** Justin Chen, Amanda Zaman, Bharat Ramakrishna, OpenBiome Team, Scott W. Olesen

## Abstract

**Objectives:** Fecal microbiota transplantation (FMT) is a recommended therapy for recurrent *Clostridioides difficile* infection and is being investigated as a potential therapy for dozens of microbiome-mediated indications. Stool banks centralize FMT donor screening and FMT material preparation with the goal of improving the safety, quality, convenience, and accessibility of FMT material. Although there are published consensuses on donor screening guidelines, there are few reports about the implementation of those guidelines in functioning stool banks.

**Methods:** To help inform consensus standards with data gathered from real-world settings and, in turn, to improve patient care, here we describe the general methodology used in 2018 by OpenBiome, a large stool bank, and its outputs in that year.

**Results:** In 2018, the stool bank received 7,536 stool donations from 210 donors, a daily average of 20.6 donations, and processed 4,271 of those donations into FMT preparations. The median time a screened and enrolled stool donor actively donated stool was 5.8 months. The median time between the manufacture of an FMT preparation and its shipment to a hospital or physician was 8.9 months. Half of the stool bank’s partner hospitals and physicians ordered an average of 0.75 or fewer FMT preparations per month.

**Conclusions:** Further knowledge sharing should help inform refinements of stool banking guidelines and best practices.

## INTRODUCTION

Fecal microbiota transplantation (FMT), the transfer of minimally manipulated stool and its associated microbiota from a healthy donor into the gastrointestinal tract of the patient,^1,2^ is a recommended investigational therapy for recurrent *Clostridioides difficile* infection.^3–7^ *C. difficile* infection is the most common healthcare-associated infection in the United States, with 462,000 cases and 20,500 deaths in the US in 2017.^8,9^ FMT’s reported safety profile and efficacy in preventing recurrence of *C. difficile* infection, approximately 80–90%,^10–18^ has inspired research using FMT to treat a wide range of microbiome-mediated indications.^2,19^

There are two main models supplying stool for FMT: *patient-selected donors* and *stool banking*.^20–25^

Under the patient-selected donor model, the patient or their guardian identifies their own stool donor candidate. The treating physician screens the candidate and processes the donor’s stool into an FMT preparation. The donor typically donates material for only that single patient. This approach places substantial logistical burden on the physician^26^ and creates delays between the determination that FMT is indicated and the delivery of therapy. For example, if a patient’s first candidate donor fails the screen, another must be found, who may also fail the screen, all before the patient can be treated.^27,28^ Furthermore, the patient-directed model poses certain risks. First, different practitioners may use variable screening standards, potentially exposing the patient to substandard screening. Second, the donor stool may be processed into an FMT treatment in an uncontrolled, *ad hoc* workspace, like a physician’s office, increasing the risk of contamination. Barriers to prompt access to FMT have also been reported as reasons for patients to seek “do-it-yourself” (DIY) FMT, which comes with significant risk to patient safety.^29^ Stool banks address many of the logistical limitations inherent to the patient-selected donor model. A stool bank is a centralized facility that screens donors, processes stool, stores FMT preparations, fulfills clinicians’ and researchers’ requests for those preparations, and monitors the safety and efficacy of the material.^22,28^ Centralized donor screening enables more rigorous and consistent safety standards, with donor qualification rates as low as 2.5%.^30^ Centralized processing is more cost-efficient and controlled: qualified donors provide material that can be used to treat many patients, and a purpose-built facility allows for stringent manufacturing quality standards.^23,25,31^ Centralized distribution minimizes the delay in treating the patient, as physicians can access a well-screened, quality-assured FMT preparation delivered overnight.

Centralized safety reporting may also contribute to improved FMT safety through a better understanding of the risks associated with human-derived microbial therapeutics and subsequent implementation of improved screening and manufacturing processes.

Although there are differences between stool banks, in part due to variation in FMT regulation between countries,^32^ stool banks generally adhere to a common-six part methodology, shown below.^5,21,23–25,31,33,34^ In some banks, these six elements are joined together by rigorous quality systems.^23,25^

1. Donor recruitment: Encourage candidate donors to undergo evaluation
2. Donor evaluation: Assess whether candidate donors qualify to donate stool
3. Manufacturing: Process donated stool into formulations suitable for use in FMT
4. Health monitoring and release: Confirm donor health before releasing FMT material from quarantine
5. Fulfillment: Provide FMT material to clinicians and researchers, and track that material
6. Patient Safety: Evaluate FMT safety and quality, and respond to emerging safety and quality issues

Despite the field’s coalescence around this overall methodology, stool banking continues to evolve, delivering material that is more rigorously screened and more carefully prepared, with greater efficiency and reliability. Because consensus guidelines for stool bank operations have been strongly informed by the practice of existing stool banks,^20,31,34,35^ improved sharing of different banks’ methodologies,^36^ exemplified by recent reports from English and Danish stool banks that emphasized the importance of quality management systems,^23,25^ should help advance consensus standards and, in turn, to improve patient care. Improved sharing will also support the development of other stool banks and invite broader participation in the improvement of stool banking methods.

Here we report on the methodology used in 2018 by OpenBiome (Cambridge, MA, USA), a large stool bank, to fulfill clinicians’ requests for FMT material to treat recurrent *C. difficile* infection under enforcement discretion.^37^ Since its founding in 2013, this bank has shipped more than 56,000 FMT preparations to a network of 1,250 healthcare facilities. We describe the bank’s overall methodology in 2018, the most recent complete year when this study began, and the bank’s outputs in that year.

## METHODS

The stool bank follows phase-appropriate current good manufacturing practices (cGMP),^38^ which includes well-defined procedures, a highly-controlled manufacturing environment, and meticulous record-keeping.

In 2018, the bank used the six-part methodology described above and outlined previously.^24^ This methodology does not necessarily reflect the bank’s current operation as changes are continuously being made to stool bank operations based on the latest scientific evidence and under enforcement discretion as well as requests for material for research purposes. Some regulatory guidance. The bank fulfills requests for FMT material to treat *C. difficile* infection under enforcement discretion as well as requests for material for research purposes. Some procedures differ between enforcement discretion and research. For clarity, only the enforcement discretion procedures are described here.

103 104

### 1. Donor recruitment

The bank encourages members of the public between 18 and 50 years old to enroll as stool donors using conventional press and social media campaigns. Because the bank requires that donated stool be passed and donated inside the stool bank’s facility, rather than at home, the campaigns only target the metropolitan area where the stool bank is located. Individuals interested in becoming stool donors complete an initial, online health questionnaire focused on eligibility criteria that commonly leads to *a priori* exclusion. Important categories used for preliminary screening include logistics (e.g., ability to donate at the stool bank at least three times per week), infectious risk (e.g., high risk travel history),^39^ and compromised microbial diversity (e.g., recent antibiotic use).^30^ Candidates who meet eligibility on the pre-screen questionnaire are invited for an on-site evaluation.

### 2. Donor evaluation

During on-site evaluation, candidates provide informed consent and sign an affidavit attesting that the health information they provide is true and complete.^40^ Next, candidates complete an in-depth donor health questionnaire, which includes questions about gastrointestinal comorbidities, metabolic conditions, neuro-psychiatric comorbidities, infectious diseases, autoimmune diseases, atopy, asthma and allergies, malignancy (e.g., colorectal cancer), surgeries or other medical history, current symptoms and behaviors (e.g., bowel habit), medications (e.g., antimicrobial therapy), diet, social history, and family history. The screening criteria were outlined by this group in more detail previously.^30^ A clinical staff member then meets with the candidate’s body temperature, blood pressure, heart rate, respiratory rate, body mass index, candidate to review and clarify their answers. Next, the clinical staff member measures the candidate’s body temperature, blood pressure, heart rate, respiratory rate, body mass index, and waist circumference. Finally, a supervising physician reviews the questionnaire and vital signs.

Candidates who pass the in-person clinical assessment undergo a three-part laboratory screening: blood, stool, and nasal swab. Samples are sent to external laboratories for testing(Table 1), including tests suggested by the European FMT Workgroup guidance and international consensus recommendations.^31,41^ Any abnormality is reviewed by the bank’s supervising clinician. Candidates who pass the laboratory screens are accepted into the stool donation program. Candidates who fail the laboratory screens are either temporarily deferred or permanently excluded from the program. For example, if any screened multi-drug resistant organism is detected, the candidate is permanently excluded as a stool donor, but a patient carrying a transient enteric pathogen may be invited to re-screen after a temporary deferral.

Candidates and donors are informed of any significant incidental findings observed during the health evaluation and monitoring process and referred to the appropriate healthcare services.

To protect the security and quality of the donor program, candidates are generally not informed about the reason for their deferral or exclusion other than to share significant incidental findings.

**Table 1:** Donor evaluation, including clinical assessment and laboratory screenings, used by the bank in 2018. This list does not reflect the bank’s current screening.*

| **Clinical Assessment** |
| --- |
| **Infectious risk factors** |
| Known HIV or viral hepatitis exposures |
| High risk sexual behaviors |
| Tattoo or body piercing within previous 6 months |
| Known history of infectious disease |
| Travel history to endemic regions with a high risk acquiring infectious pathogens |
| Risk factors for multi-drug resistant organisms (MDROs) including work in clinical environment or long-term care facility, recent hospitalization, or recent discharge from a long term care facility |
| **Potentially microbiome-mediated conditions and factors** |
| Gastrointestinal conditions (e.g., history of inflammatory bowel disease, irritable bowel syndrome, chronic constipation, chronic diarrhea, celiac disease) |
| Atopic conditions (e.g., asthma, atopic dermatitis, eosinophilic disorders of the gastrointestinal tract) |
| Autoimmune conditions |
| Chronic pain syndromes |
| Metabolic conditions (i.e., clinician assessment of height, weight, and waist circumference) |
| Neurological conditions |
| Psychiatric conditions |
| Malignancy history |
| Surgeries / Other medical history |
| Current symptoms |
| Medications including antibiotics, antifungals, antivirals, and immunosuppressants |
| Diet |
| Family history (e.g., family history of inflammatory bowel disease or colon cancer) |
| **Laboratory Testing** |
| **Serological Testing** |
| Complete blood count with differential |
| Hepatic function panel (AST, ALT, ALP, bilirubin, albumin) |
| HIV-1/2 antigen and antibodies (fourth-generation test) |
| Hepatitis A (IgM) |
| Hepatitis B panel (HBsAg, HBsAb, HBcAb) |
| Hepatitis C (antibody) |
| *Treponema pallidum* (cascade with reflex to RPR) |
| Human T-lymphotropic virus I and II (antibody) |
| *Strongyloides* (IgG) |
| **Stool Testing** |
| *Clostridioides difficile* toxin B (PCR) |
| Enteric pathogens including *Salmonella*, *Shigella*, *Campylobacter*, and *Vibrio* (culture) |
| Shiga toxin (EIA, with reflex to *E. coli* O157 culture)<sup>†</sup> |
| *Helicobacter pylori* (EIA) |
| Ova and parasites |
| *Giardia lamblia* (EIA and microscopy) |
| *Cryptosporidium* (EIA) |
| *Cyclospora* and *Isospora* (microscopy) |
| *Microsporidia* (microscopy) |
| Rotavirus (EIA) |
| Norovirus (real-time PCR) |
| Adenovirus (EIA) |
| Vancomycin-resistant *Enterococcus* (culture) |
| Extended spectrum beta-lactamase producing Enterobacteriaceae (culture) |
| Carbapenem-resistant Enterobacteriaceae (culture) |
| **Nasal Swab Culture** |
| Methicillin-resistant *Staphylococcus aureus* (culture) |

AST: aspartate transaminase. ALT: alanine transaminase. ALP: alkaline phosphatase. HBsAg: hepatitis B surface antigen. HBsAb: hepatitis B surface antibody. HBcAb: hepatitis B core antibody. RPR: rapid plasma reagin. EIA: enzyme immunoassay. PCR: polymerase chain reaction. \* Screening for SARS-CoV-2 via nasopharyngeal swab PCR was implemented in March 2020, but material donated from December 2019 and onward remains in quarantine pending regulatory review. † EIA for Shiga toxin was replaced by *stx1/2* PCR in March 2020.

### 3. Manufacturing

Donors provide donations by visiting the stool bank’s collection facility, where they are given a stool collection kit. After passing the stool on site, donors close the container’s lid and place it in a resealable plastic bag for secondary containment. A staff member labels the donation with a donor identification number and the time of passage. The donor’s health status is re-assessed at each donation as described below. Donors are remunerated for the time and travel required to provide each processed donation.

The donation is transferred to a dedicated biosafety cabinet that is cleaned with a sporicidal agent and ethanol. A trained technician opens the container and evaluates the stool for any visible pathology (e.g., contamination, discoloration, blood, or mucus). Donations with poor consistency (Bristol stool scale outside 3–5) or visual pathology are destroyed, and clinical staff are notified of the abnormality. Donations that do not meet a minimum weight of 55 grams are not cost-effective to process and are destroyed. After visual inspection and weighing, the stool is transferred to a sterile 330μm filter bag, diluted in a sterile, US Pharmacopeia-grade glycero-saline solution (12.5% glycerol in 0.90% w/v NaCl in water), and fully homogenized while still in the filter bag using a paddle blender for at least 180 seconds. Fibrous material remains on one side of the filter while bacteria, small molecules, and water are pressed to the other side of the filter.

The filtrate is diluted and aliquoted depending on the final FMT preparation. Each FMT preparation is derived from a single donor; filtrates from different donors are never mixed. All donations are processed within six hours of the donor’s initial passage, and material that cannot be processed in time is destroyed.^42^

The bank produces two liquid preparations and one capsule preparation. The first liquid preparation is 250 mL diluted at a 10:1 ratio (i.e., approximately 22.7 g of stool per preparation) and is intended for delivery by colonoscopy, sigmoidoscopy, or enema delivery. The second liquid preparation is 30 mL diluted at a 5:2 ratio (i.e., approximately 8.6 g of stool) and is intended for delivery by esophagogastroduodenoscopy or nasoenteric tube. Liquid FMT preparations are transferred to sterile polyethylene terephthalate bottles using sterile, disposable serological pipettes.

The capsule formulation is designed to be swallowed and resist degradation from stomach acid until reaching the small intestine.^43^ Each capsule (size 00) consists of approximately 275 mg of stool. The stool, combined with a lipid buffer and a glycerol buffer, is contained within an inner gelatin capsule and an outer acid-resistant shell. A capsule dose consists of 30 capsules a 30 (approximately 8.25 g of stool). From each donation that is processed into an FMT preparation, a 30 mL safety aliquot is set aside for safety testing and quality purposes. Additional aliquots are retained for safety testing or research purposes. All liquid and capsule preparations are sealed with tamper-evident bottle caps and stored at –80 °C.

To support traceability, each FMT preparation and aliquot is labeled with a barcode that links it to the donation. All steps in the manufacturing process are monitored and logged according to cGMP standards. Completed FMT preparations are kept in quarantine until released, as described below.

### 4. Health monitoring and material release

A donor’s health and eligibility for continued donation are continually assessed by five mechanisms.

First, donors must report travel or change in health status while actively part of the program, including fever, cough, congestion, change in bowel habit, nausea, vomiting, seasonal allergies, medical or dental procedures, bodily injury, and use of oral or topical over-the-counter medications. If an illness or travel event is reported, the donor is further evaluated by a member of the clinical staff, who determines if the donor should be temporarily deferred or permanently excluded from the stool donation program. For example, travel to certain countries entails a high risk for acquiring antibiotic-resistant bacteria and may lead to temporary deferral or permanent exclusion. Any donor who is temporarily deferred undergoes a partial or complete re-screening before being re-admitted to the program.

Second, at each stool donation, donors complete a short heath questionnaire. If the donor reports a change in health status or a clinical staff member’s direct observation of the donor leads them to suspect a change in the donor’s health status, a clinician interviews the donor to gather additional information. Clinical staff may determine that the donation should be destroyed, and the donor may be temporarily deferred or permanently excluded. If a donor is temporarily deferred because of a transient illness, the donor must pass a partial or complete re-screening before being re-admitted to the program.

Third, if a manufacturing technician observes a suspected stool pathology in the donation as described above, the technician takes a photograph of that donation, destroys the donation, and shares the photograph with the clinical staff. Clinical staff review the photograph and follow up with the donor to determine if the abnormal stool pathology is related to a risk of infection or an underlying disease.

Fourth, donors must agree to undergo random health checks. A clinician examines the donor’s vital signs and assesses the donor’s health status, with a focus on bowel habits, infectious risk factors, and new behaviors that may impact the microbiome. If the clinician detects any clinical concerns, the donor may be temporarily deferred or permanently excluded. Any donor who is temporarily deferred undergoes a partial or complete re-screening before being re-admitted to the program.

Finally, every donor repeats the complete set of clinical and laboratory assessments, the same set that they passed when first enrolling as a donor, approximately every 60 days. These re-assessments “bookend” 60-day collection periods. FMT preparations produced from stool donations during a collection period are released from quarantine only after the donor passes assessments “bookend” 60-day collection periods. FMT preparations produced from stool the second full assessment at the end of the collection period. It is not the case that every donation is screened. Instead, the assessments of a donor’s health at the beginning and end of a 60-day period and a review of all clinical data collected during the collection window are taken as sufficient evidence that the intervening donations are fit for use as FMT preparations.^24,44^

Individuals infected with HIV or other pathogens may not test positive until some days after their initial infection.^45^ To account for the possibility that a donor acquires a pathogen with an extended seroconversion time, FMT material release is offset from reassessments: FMT preparations produced from stool donated less than 21 days before the reassessment are only released if the donor passes the assessment at the end of the following 60-day collection period.

Material is released from quarantine only after a full review of all clinical and laboratory data in each collection period and requires approval from two clinicians and one quality assurance staff member. All material releases are performed in accordance with cGMP standards.

If a donor newly tests positive for any infectious pathogens or other clinically significant abnormalities indicating there may be a potential underlying exclusionary medical condition, all FMT preparations produced from the corresponding collection period are destroyed. Donors who newly test positive for certain pathogens, including *C. difficile* as well as chronic infections like hepatitis B and HIV, are permanently excluded from the donor program and all material collected since the donor’s last screening is destroyed. In other cases, such as rotavirus, infections are transient, and donors are temporarily deferred. As stated above, temporarily the program. deferred donors must undergo a partial or complete re-screening before being re-admitted to the program.

### 5. Fulfillment

The stool bank provides FMT preparations to gastroenterologists and infectious disease physicians treating patients with recurrent *C. difficile* infection not responsive to standard therapies.^37^ The bank also fulfills requests for FMT material for research purposes, but procedures for fulfillment and patient safety monitoring in that case differ. For clarity, only the enforcement discretion procedures are described here.

The stool bank’s website provides information on FMT regulations in the United States, description of FMT treatment modalities, and clinical guidance, as well as registration forms that interested facilities must complete and submit in order to receive material from the bank.

Institutional shipping and billing information, as well as contact information for material control, the overseeing physician, and adverse event reporting, are collected during registration. When registered partners submit a written purchase order, FMT preparations are removed from storage at –80 °C and shipped via overnight air on dry ice in Styrofoam containers. Each container is shipped with a temperature indicator verifying that the preparations remained frozen during shipping.

Registered physicians or healthcare facilities who received stool bank material submit three kinds of information back to the stool bank. First, they must complete a log confirming that each FMT preparation was frozen upon arrival and report whether it remains in inventory, was used in treatment, or destroyed due to expiration or other reasons. Second, they are asked to complete a clinical follow-up form for each patient within 8 weeks of the FMT procedure, indicating the severity and subtype of *C. difficile* infection that was treated and the patient’s outcome following FMT. Finally, they are contractually required to report any serious adverse event (defined as death, life-threatening health event, hospitalization, disability or permanent damage, congenital event. anomaly, or other serious important medical event) to the stool bank within 24 hours of the event.

### 6. Patient Safety

When a clinician reports a serious adverse event to the bank, the bank begins an investigation. Safety staff appraise the event with respect to seriousness, severity, expectedness, and relatedness. Safety staff engage subject matter experts, including the stool bank’s clinical advisory board, which is an independent group of gastroenterologists, infectious diseases specialists, and other subject matter experts.

Barcoding, tracking, and quality assurance systems allow the bank to identify the donation and donor associated with an adverse event. If the adverse event presents a risk to other individuals who would receive material from the same donor, material from that donor is immediately placed in quarantine, and no more is shipped until safety and quality staff have determined that it is safe to do so. If the event involves an infectious pathogen, safety and quality staff can retrieve the safety aliquot taken from that donation and test it for the presence of the pathogen.

Depending on the investigation’s findings, material from the donor may be destroyed or recalled. The donor may be permanently excluded from the donation program.

Findings from the investigation are entered into a safety database with MedDRA terminology. (MedDRA, the Medical Dictionary for Regulatory Activities terminology, is the international medical terminology developed under the auspices of the International Council for Harmonisation of Technical Requirements for Pharmaceuticals for Human Use.) Data are analyzed through an analysis of similar events to detect safety signals which may indicate a trend or safety risk. Serious adverse events considered related to the FMT are reported to the appropriate regulatory authorities through the Council for International Organizations of Medical Sciences or the MedWatch^46^ system. Regular reviews of safety data inform additional safety practices and risk management strategies in a continuous improvement process.

## RESULTS

Between March 2018 and July 2018, 731 candidate stool donors completed a survey asking for their motivations for donating stool. Most donors’ motivations include helping *C. difficile* infection patients, advancing research, and receiving the per-donation compensation (Figure 1). Donors who pass screenings and enroll in the program remain for varying amounts of time: more than half of donors who donated in 2018 were active donors for less than 6 months, but one donor who donated in 2018 had been active for 3.4 years (Figure 2).

**Figure 1:**
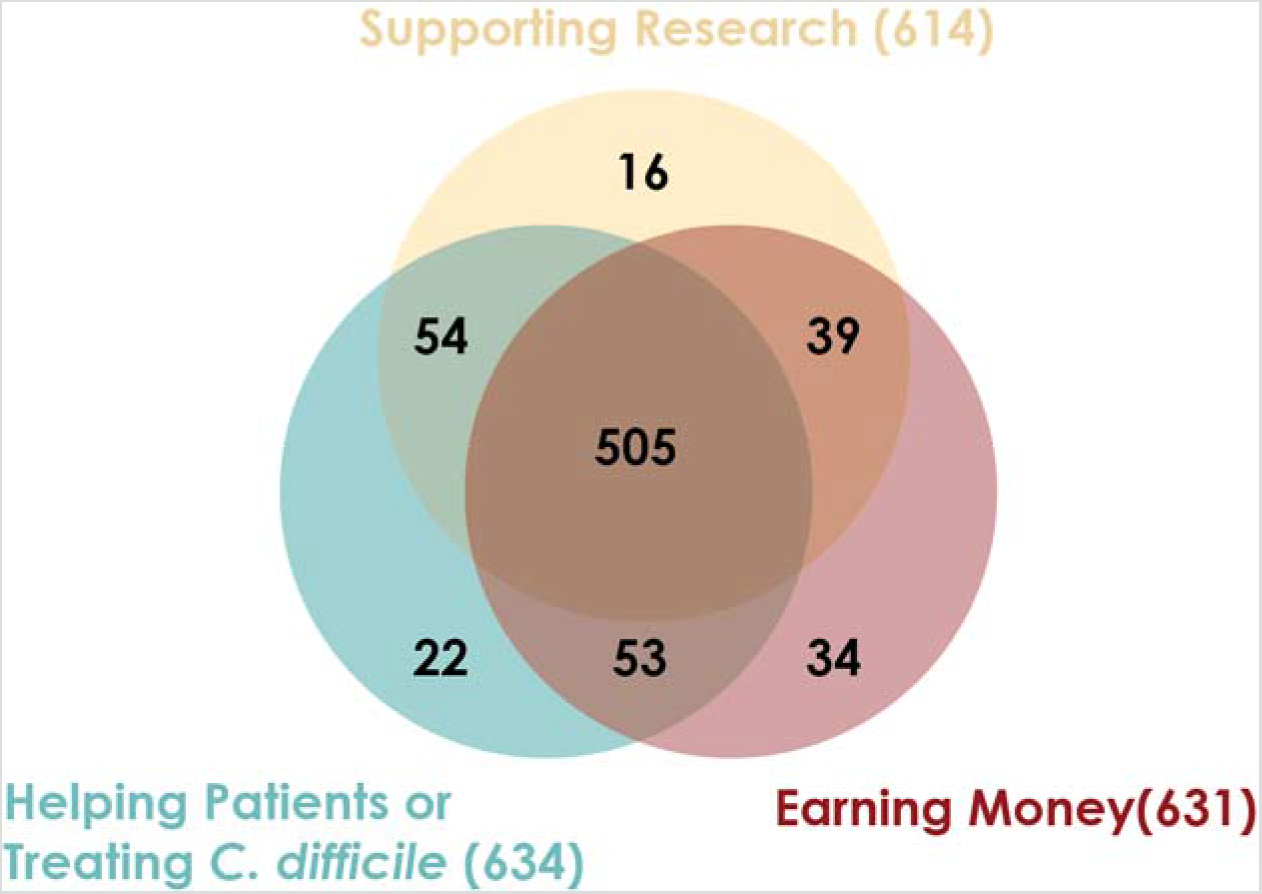
Stool donors have multiple motivations for providing stool. Between March 2018 and July 2018, 731 candidate stool donors completed a survey asking their motivations for donating stool. The survey instructed candidates to indicate all motivating factors that applied to them.

**Figure 2:**
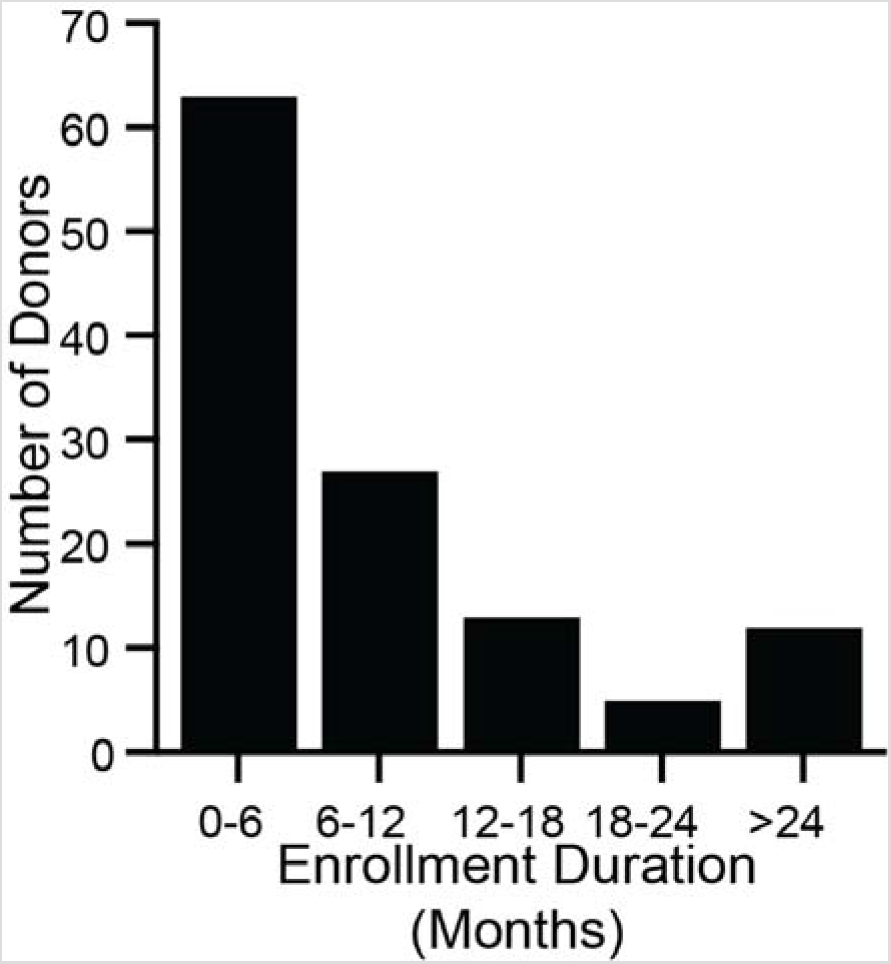
Stool donors are active over a wide range of time. During 2018, 120 donors made at least 1 donation that was processed into an FMT preparation. For each of those donors, enrollment duration was calculated as the time between a donor’s first donation (which may have taken place before 2018) and their last donation in 2018. The minimum enrollment duration was 21 days, and the maximum was 3.4 years. The median enrollment duration was 5.8 months (interquartile range 3.2 months to 12.1 months).

In 2018, the stool bank received 7,536 donations (Figure 3A) from 210 donors (Figure 3B), an average of 20.6 donations per day. 7% of donations (516/7,536) were used for stool screening purposes, to assess the donors’ health. 36% of donations (2,749/7,536) were rejected due to the donation’s low weight, poor Bristol stool score, visual stool pathology, or because the donation could not be processed within 6 hours of passage. The remaining 57% of donations (4,271/7,536) were processed into liquid or capsule FMT preparations (Figure 3A). Donors varied in their productivity and the proportion of their samples that were rejected or processed (Figure 3B). The time between a donation’s passage and when it was processed into an FMT formulation varied but was always less than 6 hours (Figure 3C).

**Figure 3:**
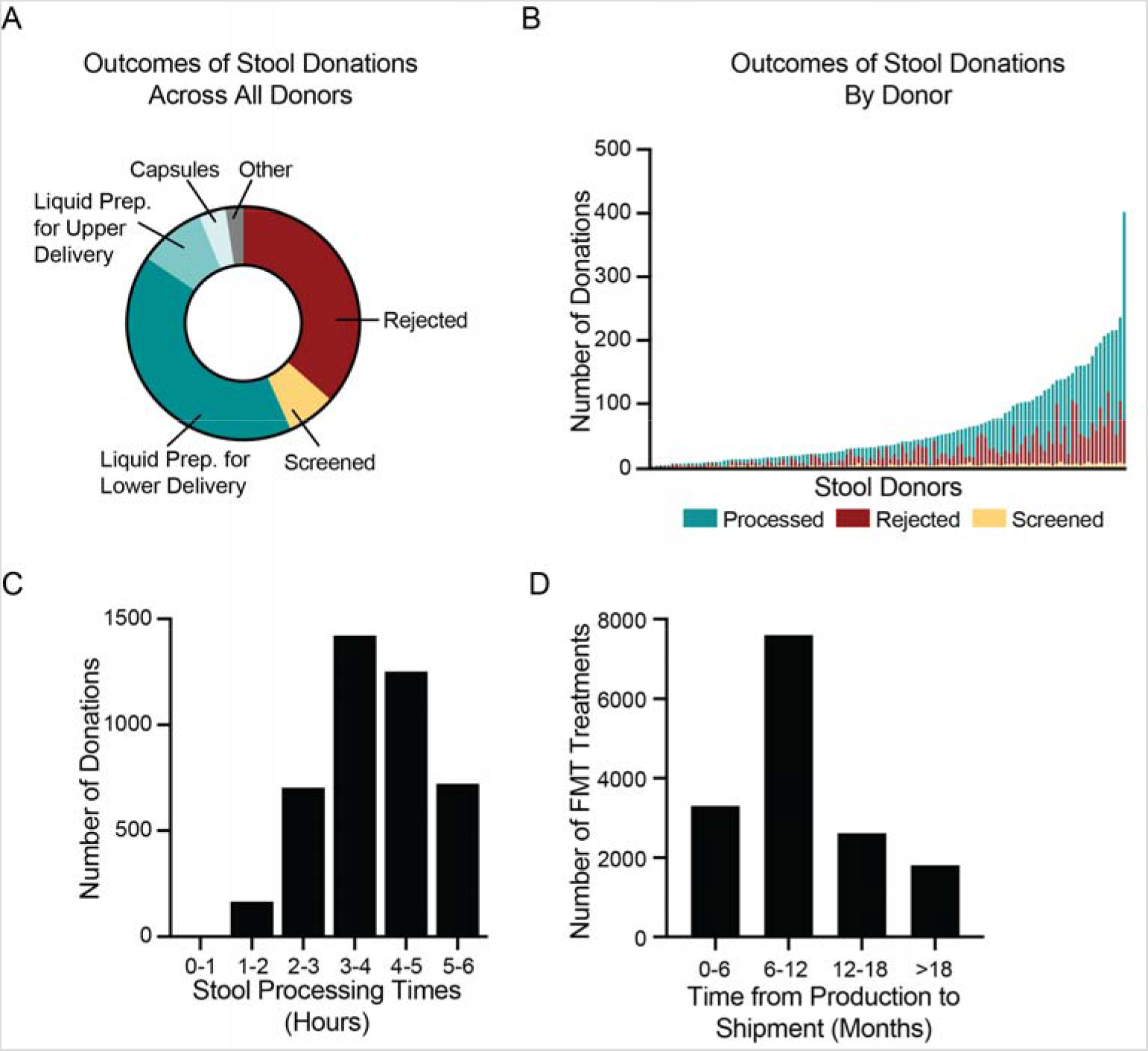
Donated stool is inspected for quality, processed into FMT treatments, and shipped to hospitals and physicians. (A) In 2018, the bank received 7,536 donations. 2,749 donations (36%) were rejected, due to visual detection of potentially pathological morphology, low weights, or failure to process the stool within six hours of passage. 516 donations (7%) were used for screening purposes to assess the health of donors. Of the remaining 4,271 donations (57%), 3,099 were processed into lower-delivery liquid preparations (e.g., for colonoscopy), 701 into upper-delivery liquid preparations (e.g., for nasoenteric delivery), 285 into capsule preparations, and 186 into other preparations. (B) During 2018, 120 stool donors provided at least 1 stool donation that was processed into an FMT preparation. Donors varied in the number of donations as well as proportion of donations that were processed, rejected, and screened. The most productive donor made 402 donations in 2018. (C) In 2018, the bank processed 4,271 stool donations into FMT preparations. All donations were processed within six hours of passage. The fastest time to processing was 47 minutes. The longest time was 5.99 hours. The median processing time was 3.9 hours (interquartile range 3.2 to 4.7 hours). (D) By 1 July 2020, the bank had shipped 15,323 of the FMT preparations produced in 2018. Preparations were shipped between 2.2 and 28 months after production. The median time to shipment was 8.9 months (interquartile range 6.3 to 13.3 months). Because not all material produced in 2018 had been shipped as of 1 July 2020, the data are skewed towards earlier shipment.

By 1 July 2020, the bank had shipped 15,323 of the FMT preparations produced in 2018 to physicians and hospitals for clinical use. Due to the complex logistics of health screening and quarantine releases, very few FMT preparations are shipped less than 4 months after they are produced. The median time between production and fulfillment was 8.9 months (Figure 3D).

To quantify how different hospitals and physicians used different amounts of the bank’s material, we measured the number of FMT preparations shipped to each recipient in 2018, regardless of when those preparations were manufactured. In 2018, 12,453 preparations were shipped to 968 recipients (Figure 4). One recipient received 141 preparations, an average of 11.75 per month, but the median number of preparations was 9, or 0.75 per month. Thus, half of hospitals and physicians who received material from the bank treated an average of less than 1 patient per month with FMT using that material.

**Figure 4:**
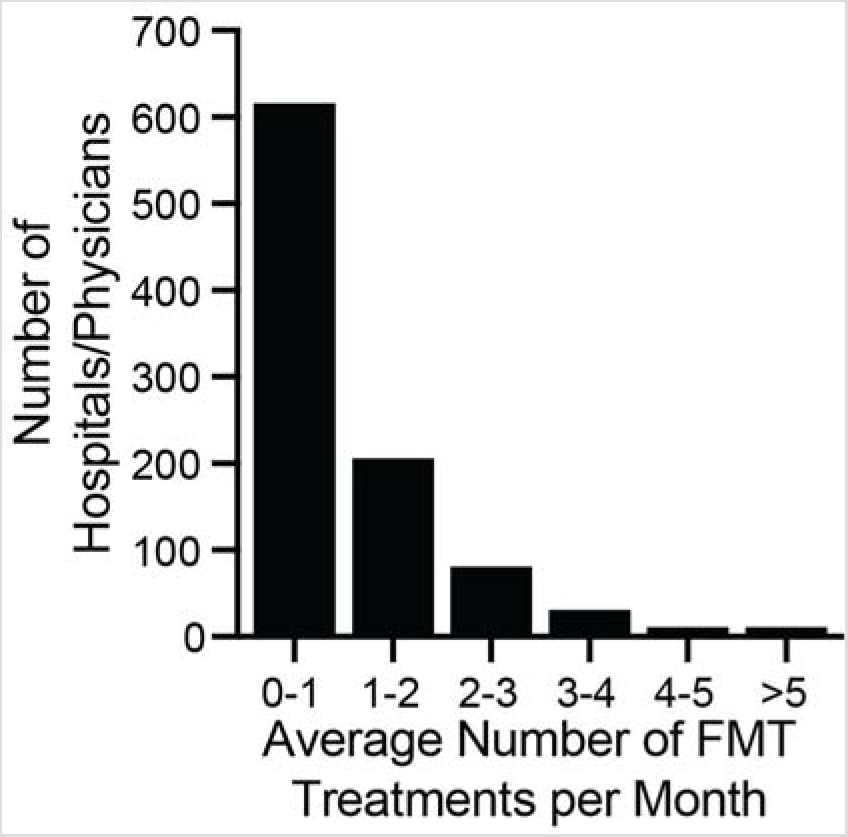
Monthly volume of preparations by physician/hospital during 2018. The number of preparations per month was computed by dividing the total number of shipped units by 12. The median number of ordered preparations across physicians/hospitals was 9 per year, or 0.75 treatments per month (interquartile range 0.25 months to 1.4 months). The maximum number of preparations ordered was 11.75 per month. The majority of physicians/hospitals ordered an average of less than 1 FMT preparation per month.

## DISCUSSION

In 2018, the stool bank’s protocol was designed principally to improve the safety and accessibility of FMT for treating *C. difficile* infection in a specific regulatory environment,^37^ using donors from a limited geographic location, and guided by the scientific and medical understanding of FMT at the time. The protocol described here would require modification if implemented in a different regulatory environment^23,32^ or geography.^47^ Furthermore, as the field’s understanding of the safety profile and molecular mechanisms of FMT improves, there will be further opportunities to optimize human-derived microbial therapies in general and stool banking protocols in particular.

We shared these methods to help inform consensus standards with data gathered from real-world settings, to support the advancement of similar operations, and to invite broader participation in the improvement of these methods. To help explore the thematic areas in which stool banking protocols could be improved and adapted, we lay out and discuss five scenarios about a hypothetical donor’s lifecycle.

### Scenario 1: A healthy donor

In this scenario, the hypothetical donor passes initial screening and subsequent health checks. Donated stool is used to prepare FMT preparations, which successfully resolve patients’ *C*. *difficile* infection. No adverse events related to this donor’s material are reported, and the donor eventually leaves the program for their own reasons. Even this scenario presents multiple questions about optimal stool banking protocols.

First, the stool bank compensates donors $40 USD to remunerate them for the time and effort associated with making a usable on-site donation. Compensation should be high enough to fairly remunerate donors for their efforts. However, compensation should also avoid perverse incentivization or coercing candidate donors into sharing private health information and human biospecimens. The optimal compensation for stool donors is an open question: while whole blood donors and organ donors in the US are not compensated, plasma donors, sperm donors, and egg donors are.^31,48–52^

Second, the requirement that donations be passed on-site limits the bank’s donor pool to one metropolitan area. Although stool from any donor passing the stool bank’s screens appears equally efficacious for treating *C. difficile* infection,^53–56^ the same may not hold true for other diseases. For example, it may or may not be important for donors and recipients to be geographically “matched” to ensure maximally safe and effective FMT.^57–59^

Third, because stool from any donor in the bank’s program appears equally efficacious, the stool bank does not direct material from particular donors to particular patients except as part of specific research protocols. However, it may at some point become clear that individual donors(“super donors”)^60^ or donations with particular characteristics (“superstool”)^61^ yield more effective FMT preparations for treating *C. difficile* infection or other indications. As the field’s understanding of FMT grows, the universal donor approach may require adjustment.

Fourth, the active ingredient of human-derived microbial therapeutics like FMT, although suspected to be live or viable bacteria, is not definitively known,^62^ which makes the optimal manufacturing protocol unclear. Current manufacturing processes, such as aerobic preparation and freezing FMT preparations, are supported by clinical evidence^63–65^ but may require adjustment as clinical and scientific understanding of FMT grows. Furthermore, a human-derived therapeutic’s active ingredient may not be the same for all theorized indications.^66^ Current testing for FMT potency focuses on donations’ viable bacterial density,^67^ but there is the possibility that future research into the variability of human stool and the correlation between those variations and the efficacy of FMT for treating various indications may reveal a more accurate predictor of FMT potency.^61^

Fifth, even with rigorous health screenings, human-derived microbial therapeutics present risks that patients should be counselled on during the informed consent process. Screening standards used by the bank for FMT material are informed by criteria set forth by regulatory bodies, blood banks, an independent clinical advisory board, national and international stool banking consensus guidelines,^5,26,31,35,68^ and patient outcomes reported by partner hospitals and physicians. However, not all known pathogens are screened for, and previously unknown pathogens or other risk factors can arise in human-derived material.^69^ Thus, screening standards for human-derived microbial therapeutics require continuous re-evaluation and updating.

Finally, even if human-derived microbial therapeutic material is screened for a pathogen or condition, different test modalities may lead to different results. Ideally, a screening assay reliably determines whether a donor’s material carries the minimum infective dose of a pathogen. In practice, the fact that a donor or their material screens negative for a pathogen on reliably determines whether a donor’s material carries the minimum infective dose of a an available assay does not fully negate the risk that their donated material carries that pathogen, and patients should be counselled on the risk of acquiring pathogens during the informed consent process.

### Scenario 2: A donor fails the initial screen

In this scenario, the potential donor is excluded during the pre-screen survey, on-site evaluation by a clinical staff, or laboratory testing. The candidate never provides a stool sample. This scenario highlights two major areas of ongoing development.

First, just as blood donation regulations aim to reduce risk “to the lowest level reasonably achievable without unduly decreasing the availability of [blood]”,^70^ so stool donor screening procedures aim to limit the risk of FMT without unduly restricting access to FMT. However, the optimal screening battery remains an area of active development, and screening for every possible pathogen may not be the best approach. For example, given the high prevalence of prior exposure to Epstein-Barr virus (EBV) and cytomegalovirus (CMV) as well as the lack of reported patient events related to transmission of these viruses via FMT, international guidelines do not recommend that donors be excluded based on their exposure to these viruses.^31^ Instead, patients at risk of CMV or EBV infection should therefore be appropriately counselled on the risks, and alternatives to FMT should be considered. The label on the bank’s shipped material includes a disclaimer to this effect.

Second, the optimal screening battery may be different for different patient populations or geographies. As a hypothetical example, it may become clear that testing for some pathogen or risk factor is important for FMT safety, but only when used in a certain patient population, such as immunocompromised patients, thus requiring a tailored match between donor screening and patient population. This tailoring is practiced in blood banks, which provide, for example, CMV-negative blood to CMV-naïve patients. Similarly, stool banks in different geographies should determine their screening criteria based on the local burden of disease to design locally appropriate screening programs.

### Scenario 3: A donor is permanently excluded from the program

In this scenario, an active donor contracts an exclusionary infection, such as *C. difficile* or HIV, or is diagnosed with a disqualifying, potentially microbiome-related condition, such as hypertension or rheumatoid arthritis, for the first time. The donor is permanently excluded from donation program, and FMT preparations made from their donations since their last battery of negative screens are destroyed. The stool bank may also destroy material made before the previous negative screening.

Similar to Scenario 2, the optimal procedures for determining criteria for permanent donor exclusion are an area of active development. While HIV, hepatitis B, and hepatitis C are obvious criteria for permanent exclusion, the need to permanently exclude donors because of other infectious diseases or because of potentially microbiome-mediated conditions will continue to be refined. As the field’s understanding of the microbiome’s role in disease grows and the potential risk for transmission of microbiome-mediated diseases becomes more clear, human-derived microbial therapeutics, including FMT and stool banking procedures, will likely adapt to improve patient safety.^30,71,72^

### Scenario 4: A donor is temporarily deferred after contracting an acute infectious disease

In this scenario, a donor self-reports, tests positive for, or is diagnosed with an acute infection, such as a viral upper respiratory infection or acute gastrointestinal illness. The donor is temporarily deferred and any material from the current stool collection period is destroyed. The deferral is maintained at least until the donor is asymptomatic, usual stool patterns have temporarily deferred and any material from the current stool collection period is destroyed. The returned, and the donor passes a partial or complete re-screening, which includes tests for infectious diseases. Stool bank clinical staff use their clinical judgment to determine if the deferral should be extended to account for pathogen shedding that can continue after resolution of symptoms. Although the fecal shedding periods for some pathogens have been studied, there is no published clinical guidance about the appropriate intervals for stool donor deferrals after acute infection, making this an aspect of stool banking that requires further discussion and development.

### Scenario 5: Patient experiences an adverse event following FMT

In this scenario, a clinician treating a patient with FMT material from the stool bank under enforcement discretion reports that the patient experienced an adverse event. Centralized safety reporting and surveillance may contribute to improved FMT safety through a better understanding of the risks associated with human-derived microbial therapeutics and subsequent implementation of improved screening and manufacturing processes, but there are important areas of active development with respect to FMT safety.

First, improving methods for collection, integration, and operationalization of safety data for human-derived microbial therapeutics is an important area of ongoing development. Safety data about FMT is obtained from three main sources. Clinical trials carefully track adverse events from a relatively small number of enrolled patients. Registries like the National Pediatric FMT Registry^73^ and the American Gastroenterological Society’s FMT National Registry^74^, which aims to track 4,000 FMT patients for ten years, are critical tools in identifying and informing the mitigation of any long-term risks of FMT, an area of ongoing clinical interest.^75^ Stool banks can collect large amounts of real-world data^76^ that may be more representative of relevant patient populations than data collected from clinical trials,^77^ but improved reporting of adverse events to stool banks remains an important challenge for the field. Data from each of these 3 sources are valuable and complementary, and they should be used in concert to improve donor screening and manufacturing protocols for all human-derived microbial therapeutics.

Second, assessment of any potential causal relationship between FMT material and a single reported adverse event is often complicated by the patient’s comorbidities. For example, although several reports describe worsening inflammatory bowel disease flares after FMT, it has not yet been determined whether or not FMT contributed to those flares: patients may have experienced those flares even if they had not undergone FMT.^78^ Controlled trials and registries remain key tools to address these questions.

Third, in the case of an adverse event caused by an infection, linking pathogens in donated stool to the infective pathogen in the patient is an area of active development. Whole genome sequencing has emerged as the leading tool to assess whether FMT transferred a pathogen,^79^ but definitive proof of transmission or non-transmission via human-derived material is not always possible. In particular, retained stool samples from the donor and the patient are essential for definitively investigating a possible pathogen transmission but collecting patient samples, to support comparison with the donor samples retained by stool banks, may not yet be common practice at most facilities.^25^

## Conclusion

Optimal stool banking methods likely depend on the target patient population, evolving regulations, and emerging scientific understanding about the microbiome. Here we described and discussed the stool banking protocol used by a large stool bank in 2018 to illustrate current practice and to highlight areas for potential improvement. We hope this report is a step toward analyses that can help inform evidence-based stool banking protocols.

## Data Availability

All data underlying the analyses are available in the Supplemental Material.

## DECLARATIONS

## Data availability

All data underlying the analyses are available in the Supplemental Material.

## Authors contribution statement

JC and SO conceived the project and performed the analysis. JC, AZ, BR, and SO wrote the original draft. All authors reviewed and revised the manuscript.

## OpenBiome Team Members

(The “OpenBiome Team” includes current and former OpenBiome employees, members of OpenBiome’s Board of Directors, and members of OpenBiome’s independent, unpaid Clinical Advisory Board.) Jessica R. Allegretti, Kanchana Amaratunga, Daniel Blackler, Michael Bougas, Shrish Budree, Laura Burns, Jessica W. Crothers, Michael Dickens, Jacob Dixon, Nancy DuBois, Carolyn Edelstein, Michael Edmond, Ryan J. Elliott, Monika Fischer, Dinara Gabdrakhmanova, Richard Hunt, Stacy Kahn, Daelyn Kauffman, Deberly Kauffman, Colleen Kelly, Ciaran Kelly, Clara Kerwin, Kelly Ling, Daniel Martin, Gina Mendolia, Rodrigo Munoz, Kelsey O’Brien, Majdi Osman, Allison Perrotta, Sonny Qazi, Alexandra Scheeler, Monica Seng, Madeline Sovie, Neil Stollman, Zac Stoltzner, Elaine Vo, Kurt Warren, Karl Yoder, Charles Young

## Ethics

This study was conducted as part of study protocol 1280272 approved by New England Independent Review Board.

MedDRA® trademark is registered by IFPMA on behalf of ICH.

